# Gene–Morphology Alignment via Graph-Constrained Latent Modeling for Molecular Subtype Prediction from Histopathology in Pancreatic Cancer

**DOI:** 10.64898/2026.03.05.26347711

**Authors:** Alejandro Leyva, Abdul Rehman Akbar, Muhammad Khalid Khan Niazi

## Abstract

Molecular subtyping of cancer is traditionally defined in transcriptomic space, yet routine clinical deployment is limited by the availability and cost of sequencing. Meanwhile, histopathology captures rich morphological information that is known to correlate with molecular state but lacks a principled, mechanistic bridge to gene-level representations. We propose a graph-constrained learning framework that aligns morphology-derived signals with a fixed, data-driven gene network discovered via hierarchical Monte Carlo screening. We can derive new gene sets for classification using random sampling, and use the coexpression network of that graph to enforce the learning of a pure morphology model without using gene expression. The resulting model performs subtype prediction using morphology alone, while being explicitly forced to operate through a gene-structured latent space. Structural alignment is enforced during training. For Moffitt classification in pancreatic cancer using PANCAN and TCGA datasets, the model has a reported 85% AUC using an alternative gene set’s network structure, while the alternate gene set itself has an 84% AUC in all patients that were classified with subtyping with pancreatic cancer in the dataset. This framework demonstrates that virtual transcriptomics can provide biologically grounded molecular insights using only routine histopathology slides, potentially expanding access to precision oncology in resource-limited settings.

## 2 Introduction

Pancreatic ductal adenocarcinoma (PDAC) is the 10th most diagnosed cancer in the United States but has the third highest 5-year mortality [1]. Detecting early onsets of PDAC is difficult, which has led to the use of transcriptomic profiling for prognostic evaluation [2]. Although histopathological imaging provides a direct view of the cancer, it is limited by the region from which it was taken, as well as by the subjective perception of the evaluator [3]. In contrast, genomic information is valid across all regions of the tumor but is limited by the time at which the test was taken and does not prove whether genetic changes are due to treatment [4].

Moffitt’s basal–classical PDAC testing mechanism used non-negative matrix factorization of 50 biologically critical genes to determine two genetic subtypes of PDAC [5]. The basal subtype is characterized by poor differentiation, catabolism, and low mucinal organization; it has been shown to result in poor prognosis and lower chemosensitivity [6]. The classical subtype has been shown to be the opposite, with better prognosis, chemosensitivity, functionalization, and differentiation [7].

Today, two versions of the Moffitt system are commercially available through approval by the Clinical Laboratory Improvement Amendments (CLIA) of the United States: the Moffitt 170-gene test, which is less common, and the PURIST set [8,9]. The PURIST genetic test is based on the moffitt test and is the most commonly used version of Moffitt [10]. The test is composed of 16 of the original 50 Moffitt genes and uses pairwise sampling, while other Moffitt tests use gene enrichment or clustering, similar to other clinical tests such as PAM50 [11]. These tests have grown in demand and have been shown to be prognostic for pancreatic cancer [12].

Since genetic testing can be expensive and slow to yield results, the fields of digital pathology and oncology have developed deep learning methods to determine the genetic subtype of a patient using only histopathology imaging on Hematoxylin and Eosin-stained slides (HE) [13]. This is convenient, as biopsies are usually taken beforehand for pathological evaluation to determine tumor stage, among other variables [14]. This is done by selecting patients who have undergone biopsy and molecular testing or bulk RNA sequencing and training models to identify patterns in HE images that help determine whether a patient is classical or basal [15].

The issue with these models is that they depend on the validity of the molecular test itself and provide no direct means of identifying which image features correspond to specific gene expression patterns [16]. There is ongoing literature on how to make deep learning accessible and interpretable, and the National Institutes of Health (NIH) invested $2.3 billion in artificial intelligence, constituting 5% of the NIH budget as of 2023 [17]. In addition, Moffitt’s study is comprehensive and its prognostic capability is well established, but there remains a question as to whether other genes could serve as viable candidates or separators for testing.

This becomes a sampling problem, as over 160,000 genes are reported in bulk RNA sequencing and must be reduced to 50 genes, or ideally, just one. Non-negative matrix factorization and other optimization algorithms are effective when genes are chosen beforehand, but these algorithms are designed to impose a discrete boundary on a given sample set that depends on the applied basis function [18].

If it were possible to determine which phenotypes correspond to specific gene expression patterns and to identify alternative gene sets from a much larger pool without manual filtration, this could provide insight into how gene coexpression influences subtypes and prognosis. However, features selected by models using established methods such as attention-based multi-instance learning (ABMIL) are not always biologically correspondent and may instead reflect staining, modality, or other secondary features [19]. To bridge this gap between morphology and molecular representation, we use gene networks, where nodes represent genes and edges represent Pearson correlation (coexpression), to sample genes and model structural connectivity within tissue for molecular subtyping. A purely imaging-based model is then trained to predict whether each case is basal or classical, with a loss function that ensures adherence to the structure of the gene expression network.

## 3 Materials and Methods

To establish ground-truth labels, bulk RNA-seq samples from 180 patients from TCGA-PAAD and 617 patients from the Pancreatic Cancer Network (PANCAN), samples were matched 1:1 with a diagnostic slide to the RNA-seq data and classified into a subtype using single-sample gene enrichment analysis (ssGSEA) [20]. The total sample size was 797, with 188 high confidence cases and so on. Following the distribution of ssGSEA scores—whereby values ¡ 0 typically denote a basal-like state—samples were z-scored.

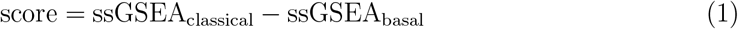

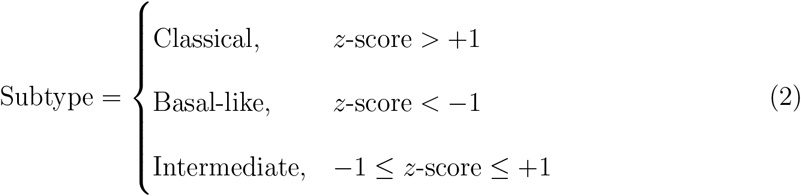

A threshold of 1 was then applied to these z-scores to categorize cases as either ‘High-Confidence Basal’ or ‘High-Confidence Classical,’ ensuring that only samples with distinct molecular profiles were used for model training.

Unprocessed bulk RNA-seq samples were indexed using GRCh38, compacted using Salmon, and converted into transcripts per million (TPM), which were then calculated and summated to produce the gene expression for each Moffitt gene in bulk RNA-seq. All computations were performed on the Ohio Supercluster on the Ascend platform with NVIDIA A100 GPUs.

To construct the graph-constrained latent space, we developed a multi-stage gene sampling workflow designed to identify the most biologically informative features from the transcriptomic data. The gene sampler has three stages, the first being preliminary filtration. If genes have no variance, no expression, or no correlation above 0.5 with at least one other gene, the gene is filtered out. Through this initial pruning, the starting set of 160,000 genes was reduced to approximately 13,000 genes. All parameters for this pruning are shown in Table 1.

**Table 1:** Key parameters for the hierarchical Monte Carlo gene selection algorithm.

| Parameter | Value | Description |
| --- | --- | --- |
| SUBSET_SIZE | 200 | Number of genes per random subset in Stage 1 |
| N_ITERS | 3000 | Maximum sampling iterations per stage and per fold |
| TARGET_AUC | 0.85 | Target AUC threshold for Stage 1 filtering |
| MIN_NON_NA | 2 | Minimum overlapping samples for gene–gene correlation |
| ABS_MIN | 0.0 | Absolute correlation cutoff for edge inclusion (disabled) |
| TOPK_EDGES | 0 | Top-$K$ edge truncation (disabled) |
| MIN_MEDIAN_EXPR | 1.0 | Minimum median gene expression threshold |
| MIN_VAR | 0.0 | Minimum variance threshold (disabled) |
| SEED | 1337 | Random seed for reproducibility |

The second stage consists of a stochastic selection process designed to identify 200-gene modules that maximize coverage across the transcriptome. Prior to selection, gene expression is normalized within each gene identity across the combined TCGA and PANCAN cohorts. This stage utilizes a Monte Carlo-inspired screening approach—performing 160–300 independent tests—to evaluate the predictive potential of random gene modules without the computational overhead of cross-fold validation.

For each 200-gene module, we establish a classification threshold based on the cohort-wide mean expression. This threshold is iteratively calibrated against the pre-classified Moffitt ground truth to evaluate the module’s ability to bifurcate samples into Basal and Classical subtypes. Modules are scored based on the accuracy of this bifurcation; those demonstrating superior alignment with the molecular ground truth are prioritized for the final network construction. Within each sample, the aggregate expression of the selected module is calculated and used to assign a preliminary subtype classification.

Since not all genes will be covered, users can alter the size of the gene module they choose. Smaller modules are computationally faster, allowing more tests, but will not cover as many genes. Larger modules will cover more genes but will be slower. This process can take between 30 minutes and about 2 hours, depending on the module size, which was optimized to be 200 genes through trial and error. A target AUC is chosen, and if any module reaches that target AUC, which in this case was 90%, the sampling stops and moves to the next stage. Otherwise, the best module is chosen and moved to the next stage. There is no guarantee that modules will have consistent accuracy, as it is improbable that the model will derive the same gene module twice in subsequent runs using a seed.

The last stage is the optimization stage within the selected 200 genes, whereby the same gene can be selected multiple times. The best 50 genes are selected from the 200-gene sample using the same network parameters shown in Table 1, and each 50-gene set is tested using cross-fold validation on the entire cohort. The best genes are then selected, and the edges between each gene within the network structure are stored as a 50×50 correlation matrix. The workflow for the sampling stages is shown in Figure1. The Laplacian of the gene matrix is taken and used as a loss function for the second component of the model. The Laplacian of the gene matrix is calculated to capture the structural topology of the co-expression network. This Laplacian is then integrated as a graph-regularized loss function for the second component of the model—a deep-learning based image classifier—ensuring that morphological feature extraction remains aligned with the underlying molecular architecture. In other words, the integration of the Laplacian regularizer proved vital in filtering out non-biological noise. Unlike traditional attention-based methods (ABMIL) which may prioritize secondary features, our graph-constrained loss function strictly limits the model’s feature extraction to patterns that align with the 50-gene signature. If two genes are strongly co-expressed in the transcriptomic space, the Laplacian forces the model to treat their morphological correlates as interdependent, thereby grounding the ‘Virtual Transcriptomics’ process in the structural reality of the tumor’s molecular landscape. The Laplacian in this case, allows for the mapping between biological dynamics between genes as a reference set by which features are selected and encoded, rather than directly integrating gene expression. As a result, genetic network structure can be used to create morphological profiles that reduce overall model loss. In this way, several genetic modules can be tested by using their network structure as a reference point, allowing for both gene discovery and spatial mapping.

**Figure 1.**
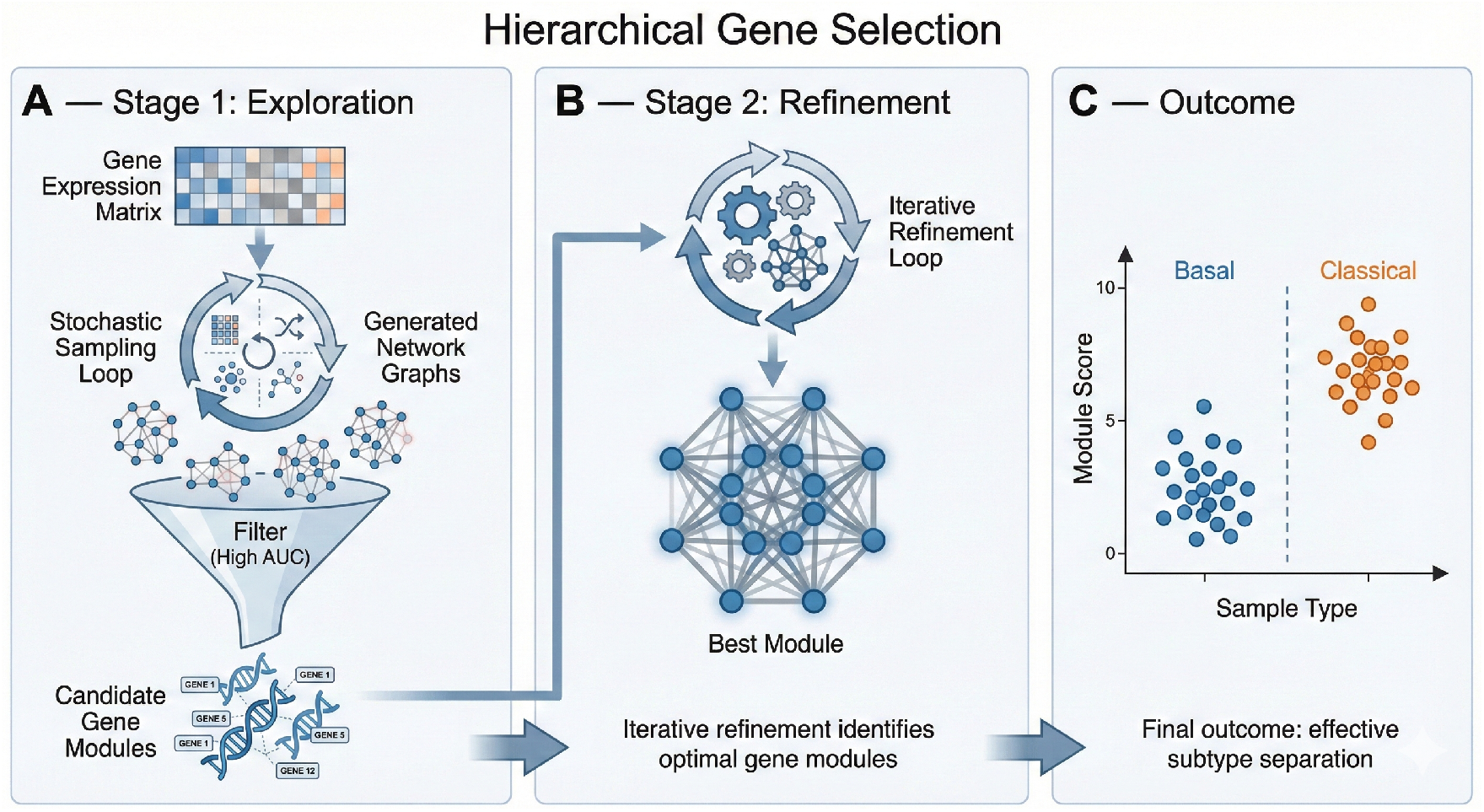
The first component of graph-constrained latent space model: Hierarchical Gene Sampling.

Gene ontology analyses were performed on the chosen gene set using R4.3.0. The morphology model utilizes 1536-dimensional UNIv2 patch embeddings extracted at 20× magnification. These patches were chosen to represent different segments of an image, and allows for the examination of morphological features that are encoded to provide better signals. Rather than mapping morphology to genetic expression that traditional spatial technologies use, this model can map genetic expression to morphology. To map these high-dimensional features to the molecular space, we employed an encoder-projection architecture: each patch embedding is first compressed into a 256-dimensional representation via a Vision Transformer (ViT)-based MLP and subsequently projected into a 50-dimensional latent feature vector. Crucially, each dimension in this latent vector corresponds to one of the 50 genes identified in the sampling stage, forcing the model to extract features with direct transcriptomic relevance. These patch-level vectors are mean-pooled to generate a patient-level representation, which is then processed through a final projection layer and a sigmoidal classifier to determine the Basal vs. Classical subtype. The model was trained using a 80/20 split across 35 epochs, optimized via a composite loss function comprising Binary Cross-Entropy (BCE), the previously described Graph Laplacian loss, and a knowledge distillation loss to align image features with the RNA-seq ground truth: Let a slide consist of *N* patch embeddings:

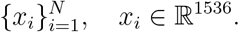

The slide-level label is:

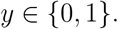

- Each patch embedding is mapped to a latent morphology space via a shared encoder:

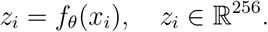
- Define a set of *G* = 50 linear projections (gene heads):

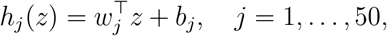

with

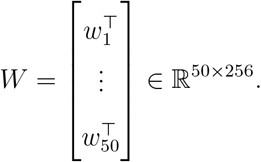

Each cell produces a 50-dimensional latent vector:

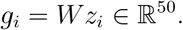

- The slide-level gene-aligned latent vector is obtained by mean pooling:

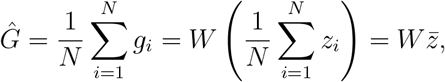

where

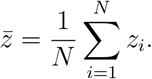
- The slide-level prediction is:

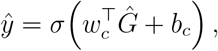

where *σ*(·) is the sigmoid function.

Let *L*_*A*_ denote the graph Laplacian of the fixed gene interaction network *A*. We enforce smoothness of the predicted gene latent representation vectors G over this graph via the regularizer

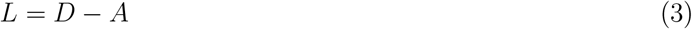

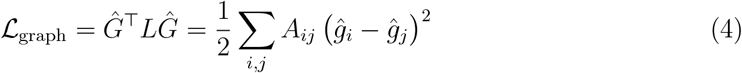

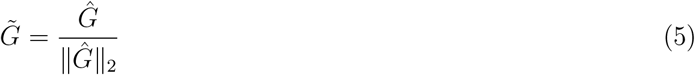

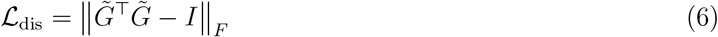

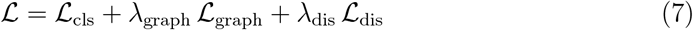

The graphical loss equation encourages gene latents connected in the molecular network to co-vary, without enforcing exact correlation matching.

To prevent degenerate solutions in which multiple gene heads collapse onto the same latent direction, we impose a competition (decorrelation) constraint, where the vector *G* is normalized in Equation 3. *F* applies the Frobenius norm. Alternative formulations include sparsity-inducing penalties or group-lasso regularization on the gene–morphology coupling weights. The last equation shows the total loss functions, composed of BCE, graphical loss, and distillation loss.

Parameters for the morphology model are shown in table 2:

**Table 2:** Configuration parameters used in the gene–morphology alignment model.

| Parameter | Value | Description |
| --- | --- | --- |
| NUM_GENES | 50 | Number of gene latent heads (per fold) |
| PATCH_DIM | 1536 | Dimensionality of UNIV2 patch embeddings |
| HIDDEN_DIM | 256 | Hidden dimension of patch encoder |
| KNN_K | 8 | Number of neighbors for spatial KNN graph |
| BATCH_SIZE | 1 | Slide-level batch size |
| EPOCHS | 30 | Training epochs per fold |
| LR | $10^{-4}$ | Adam optimizer learning rate |
| LAMBDA_GRAPH | 1.0 | Weight of gene-graph smoothness loss |
| LAMBDA_DIS | 0.1 | Weight of gene-head disentanglement loss |
| DEVICE | auto | CUDA if available, else CPU |

In summary, genes that contribute to morphological phenotypes are interconnected and can be interpreted directly, rather than using a model’s own distinction that can rely on other features.

**Figure 2.**
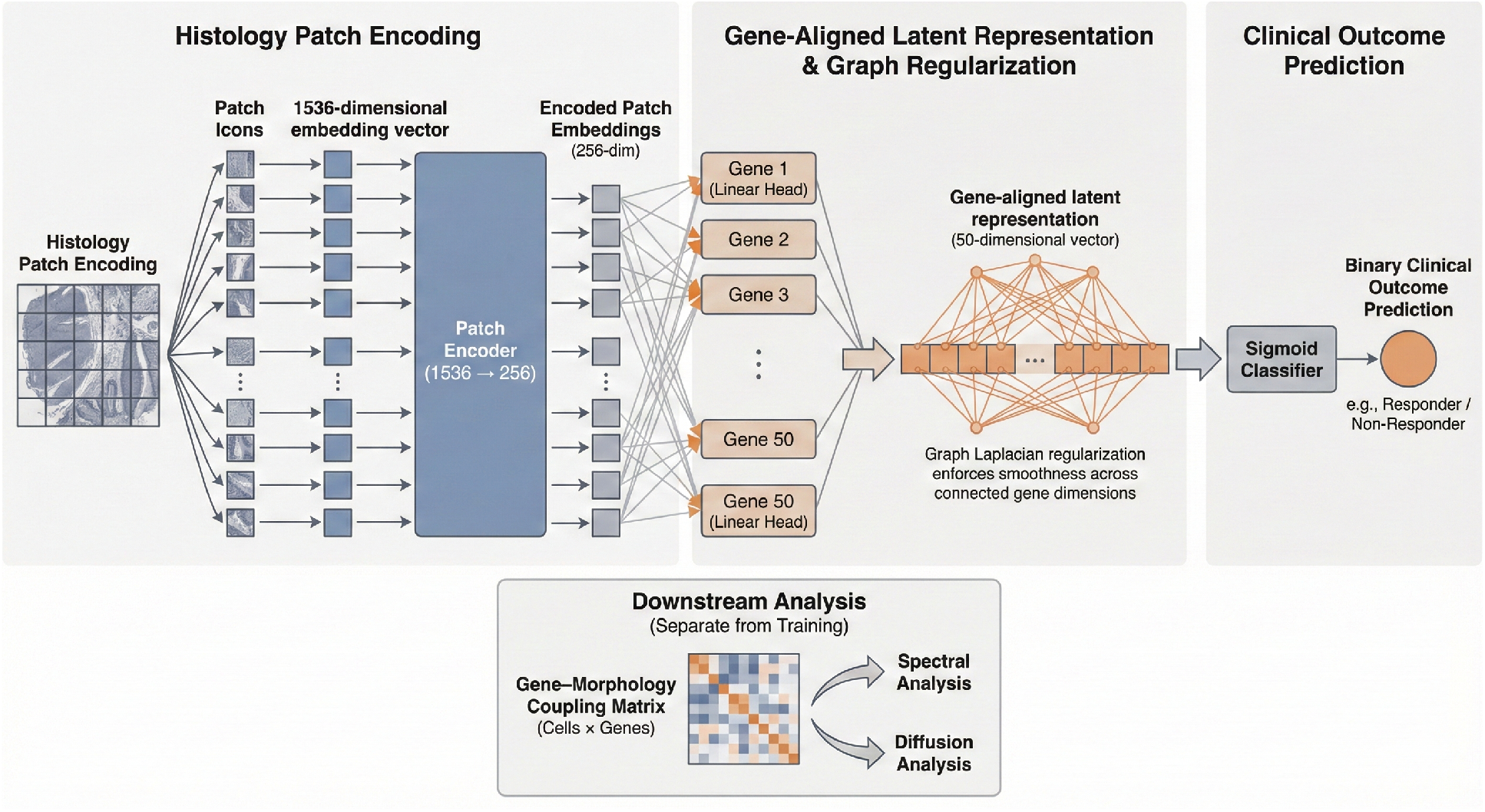
The second component of the model after hierarchical gene sampling uses the chosen gene module as a reference point, as the model learns which morphological features produce the best results based on the chosen gene set.

## 4 Results

Table 3 shows the results for the hierarchical gene sampling program, and the best optimized sample for Stage 1, which is the 200-gene sample, was tested via cross-fold validation. The 50 genes selected within that module had the best performance and were tested via cross-validation. The results show that the 200-gene sample has a lower accuracy than the 50-gene module, showing that the larger sample size contains genes with lower predictive accuracy. The module size for Stage 2 was designed to remain unchanged for the duration of the experiment. This gene sampling was tested against all samples in the cohort.

**Table 3:** Five-fold cross-validation results. Stage 2 was selected in all folds.

| Fold | $n_{\text{train}}$ | $n_{\text{test}}$ | Stage 1 AUC (Test) | Stage 2 AUC (Test) | Module Size |
| --- | --- | --- | --- | --- | --- |
| 1 | 637 | 160 | 0.793 | 0.820 | 50 |
| 2 | 637 | 160 | 0.795 | 0.819 | 50 |
| 3 | 637 | 160 | 0.754 | 0.830 | 50 |
| 4 | 638 | 159 | 0.736 | 0.854 | 50 |
| 5 | 639 | 158 | 0.788 | 0.881 | 50 |

The results for high-confidence testing (*n* = 188) are shown in table 4 with the pure morphology model, which has a decent AUC across all folds except for Fold 3. Across five-fold cross-validation, the high-confidence cohort achieved a mean validation AUC of 0.862 and mean test AUC of 0.846, with corresponding mean sensitivities of 0.720 (validation) and 0.774 (test) and mean specificities of 0.738 (validation) and 0.739 (test). In contrast, the low-confidence cohort demonstrated reduced performance, with mean validation AUC of 0.703 and mean test AUC of 0.592, accompanied by markedly lower mean sensitivities of 0.223 (validation) and 0.186 (test) despite relatively preserved specificities of 0.916 (validation) and 0.901 (test).

**Table 4:** Five-fold cross-validation performance for High- and Low-Confidence subsets. AUC = Area Under the ROC Curve; Sens = Sensitivity; Spec = Specificity.

| Fold | High Confidence |  |  |  |  |  | Low Confidence |  |  |  |  |  |
| --- | --- | --- | --- | --- | --- | --- | --- | --- | --- | --- | --- | --- |
|  | Validation |  |  | Test |  |  | Validation |  |  | Test |  |  |
|  | AUC | Sens | Spec | AUC | Sens | Spec | AUC | Sens | Spec | AUC | Sens | Spec |
| 0 | 0.895 | 0.714 | 0.933 | 0.906 | 0.947 | 0.786 | 0.630 | 0.250 | 0.882 | 0.672 | 0.238 | 0.889 |
| 1 | 0.923 | 0.941 | 0.462 | 0.853 | 0.850 | 0.625 | 0.781 | 0.000 | 1.000 | 0.629 | 0.000 | 1.000 |
| 2 | 0.769 | 0.467 | 0.867 | 0.717 | 0.600 | 0.833 | 0.660 | 0.389 | 0.795 | 0.590 | 0.333 | 0.852 |
| 3 | 0.862 | 0.714 | 0.786 | 0.935 | 0.684 | 1.000 | 0.668 | 0.000 | 1.000 | 0.431 | 0.000 | 1.000 |
| 4 | 0.861 | 0.765 | 0.643 | 0.818 | 0.789 | 0.450 | 0.776 | 0.476 | 0.905 | 0.638 | 0.360 | 0.870 |

Displayed in Figure 5 is the gene set derived from the model. Two of the genes were pseudoRNAs and were not translatable from ENSEMBL. Figure 3 shows the results from the randomly selected modules in Stage 1, as Stage 2 modules were evaluated at the fold level and averaged. Figure A shows the change in AUC over modules across the 3,000 tests, and Figure B shows the distribution centering around 0.55, as most gene modules did not show any predictive capability. Sensitivity and specificity are shown in Figure C, with no serious disproportions between gene modules across 797 samples. Figure D shows specificity against the AUC curve, demonstrating that modules with an AUC above 70% have a dramatic increase in sensitivity. Modules above 75% AUC had high specificities around 80–90%.

**Figure 3.**
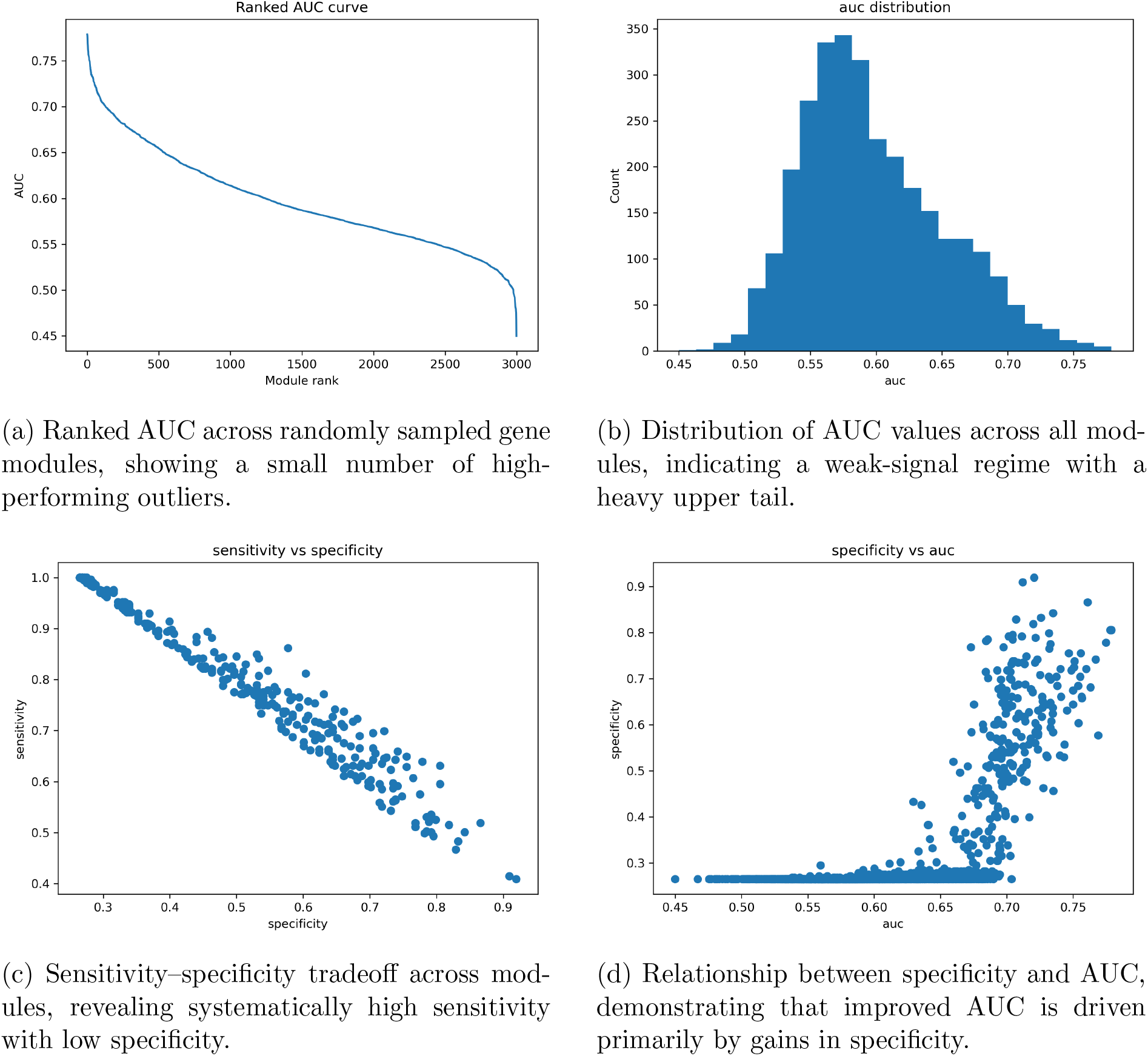
Performance landscape of randomly sampled gene modules. Ranked and distributional views of AUC establish overall signal strength, while tradeoff analyses highlight the dominance of specificity in determining discriminative performance.

**Figure 4.**
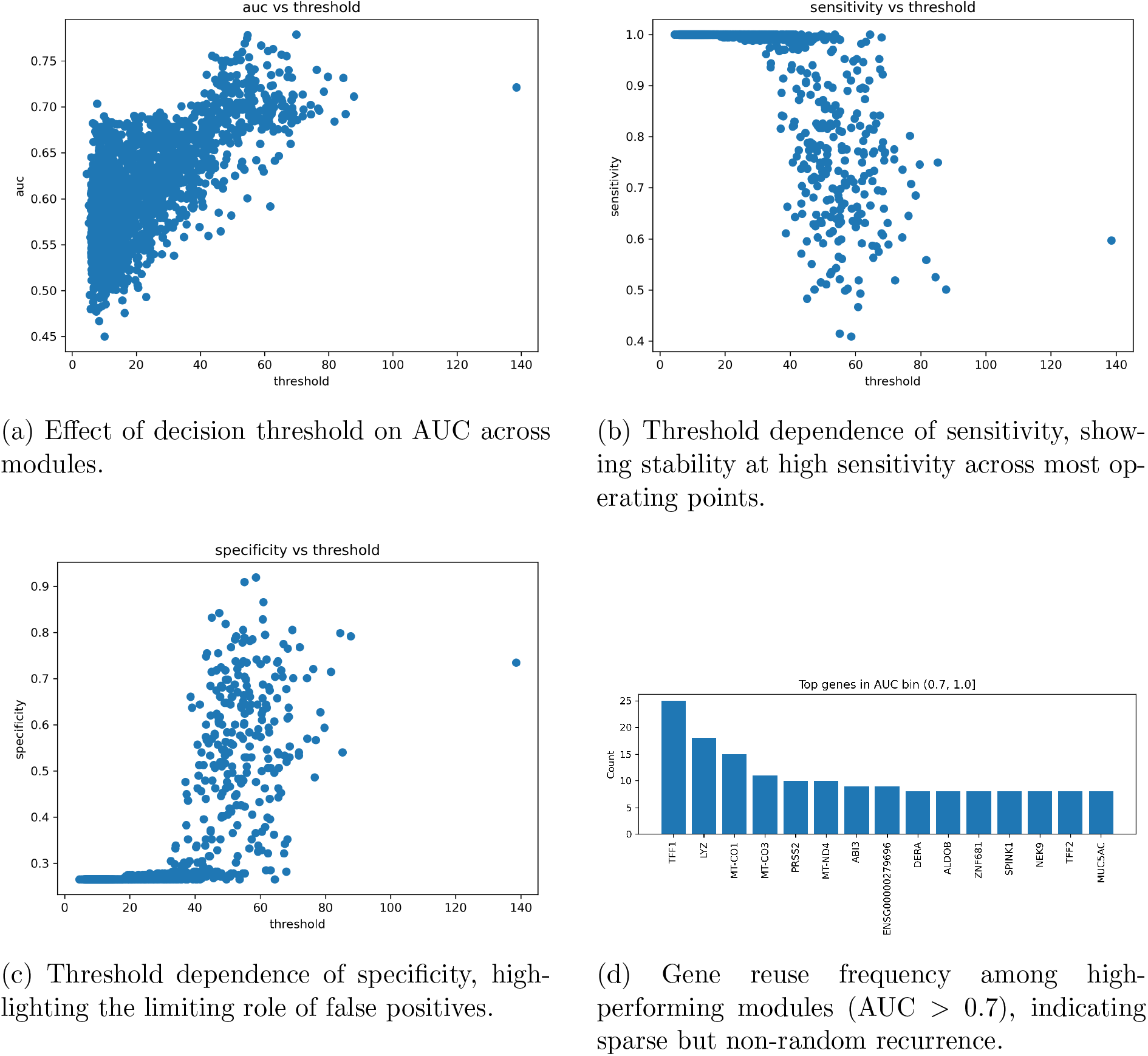
Threshold effects and gene-level signal emergence. Model performance is largely insensitive to threshold choice for sensitivity, while specificity governs overall discrimination. Gene reuse is observed only in the highest-performing modules.

**Figure 5.**
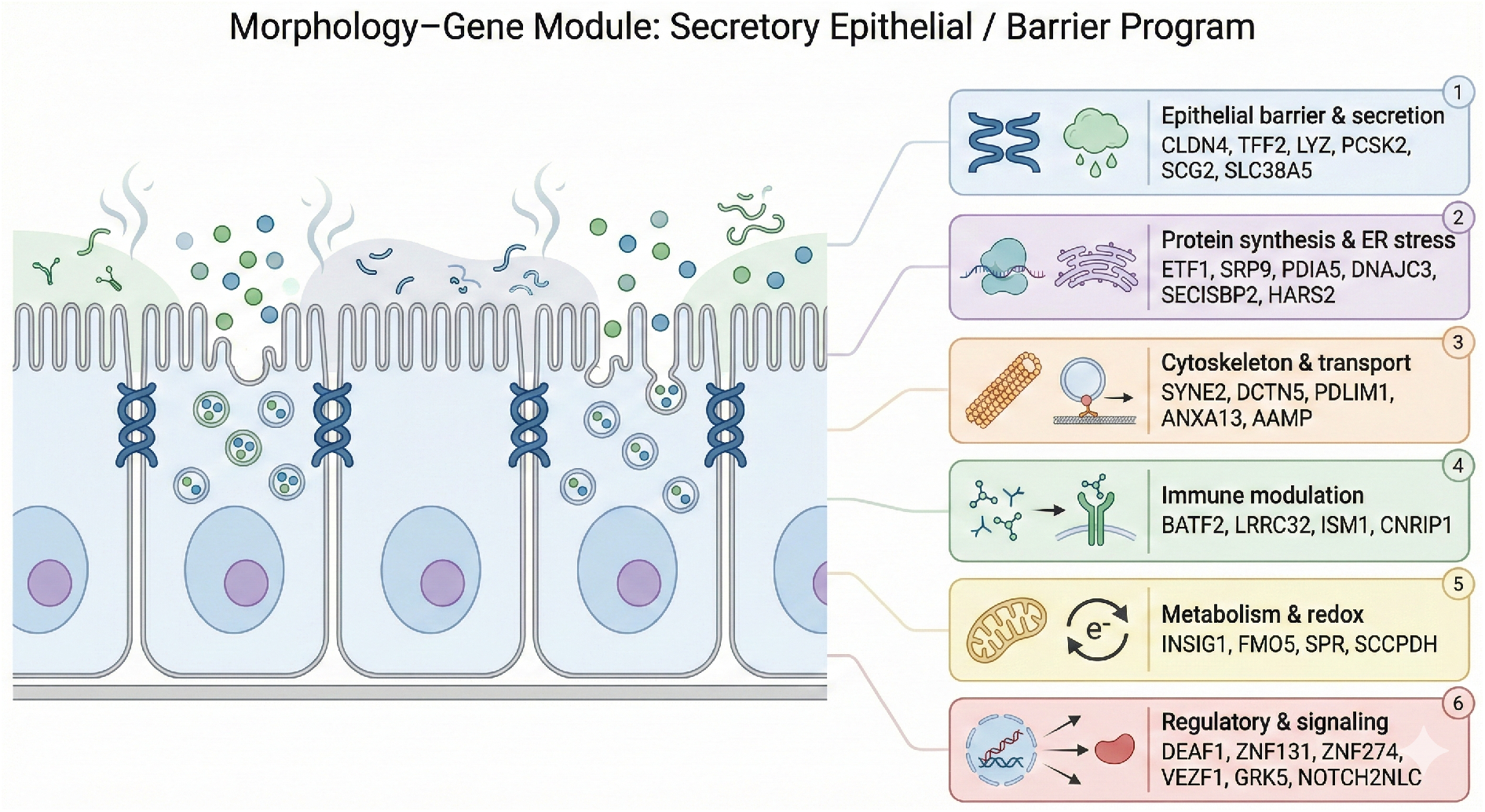
A subsample of the 50 newly derived moffitt classifiers AUC: 84%, composed of multifaceted functions and variable locations of action. Among these genes was TFF2, LYZ, and Notch2LC, which are part of the original Moffitt geneset, which had been randomly selected from the background geneset.

Figure 5 shows the validation of the threshold and recurring genes among high-AUC modules. As the threshold increased, as shown in Figure A, the AUC also increased. As shown in Figure B, sensitivity also dramatically increases as the threshold for a module reaches 60 TPM or higher, with outliers. Figure C shows the relationship between specificity and threshold, which follows the same trend as Figure B when the threshold exceeds 60 TPM. Figure D shows the top recurring genes within modules that achieved an AUC above 70%. The highest recurring genes within the highest-accuracy modules were TFF1, LYZ, MT-CO1, MTCO3, and PRSS2. The ENSG gene is a novel human locus that has not been archived by the human genome as a reference. TFF2 and SPINK1 are also recurring and are associated with the Moffitt system.

The geneset in Figure 5 characterizes 25 of the 50 new genes described, characteristically being assigned to multiple different functions, from mitochondrial, immune modulation, signaling, and protein synthesis. The full gene list is: ETF1, BSPRY, TCEA3, PDIA5, SRP9, ISM1, ZNF131, ANKRD22, PDLIM1, BATF2, RAD18, ANXA13, LRRC32, PCSK2, FMO5, SPR, DBNDD2, LYZ, SCCPDH, CLK2, PRUNE2, SMIM8, TFF2, SLC38A5, SCG2, SECISBP2, NOTCH2NLC, ZNF230, CSMD2, ZNF274, CDADC1, VEZF1, CLDN4, AGBL5, ATP9B, ENSG00000255439, ZSCAN30, DEAF1, CNRIP1, INSIG1, MKRN2OS, DCTN5, ENSG00000270953, SNORA20, AAMP, GRK5, HARS2, SYNE2, DNAJC3, RUSC2. These genes perform a variety of functions and are not clustered, but are coexpressed.

The GO analysis shown in Figure 6 includes Molecular Function, Biological Process, Cellular Component ontologies, and Gene Ontology. The two unmapped genes were not included, and because of the vast differences in function across the gene set, the probability cutoff was raised to 0.20. The top 10 genes from those analyses were used for the GO analysis and modeled. The most significant modules for the biological process analysis include the catabolism of mRNA through nonsense mutation decay, translational elongation, and protein localization to the endoplasmic reticulum. Other genes clustered to roof-of-mouth development. Amino acid transport genes were also included and labeled as statistically significant under the exploratory analysis for BP.

**Figure 6.**
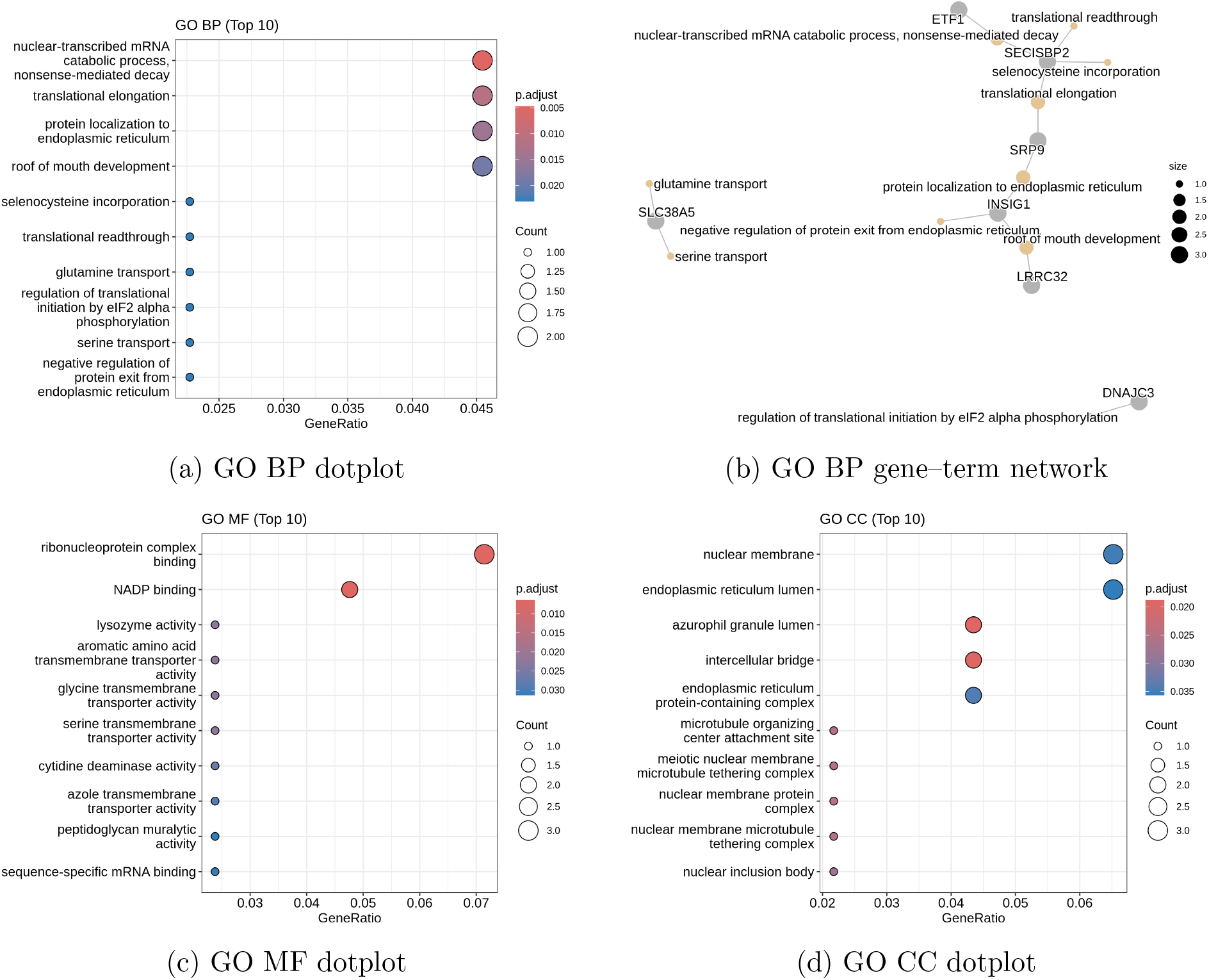
Gene Ontology enrichment analysis for the analyzed gene set. Top enriched categories are shown for Biological Process (BP), Molecular Function (MF), and Cellular Component (CC). The BP gene–term network highlights relationships between enriched biological processes and contributing genes. Only the top 10 categories per ontology are displayed.

Figure B shows the similarity network between the biological process modules, where the amino acid transport genes are separated from the main similarity network, and the main similarity network is composed of processes surrounding metabolism and catabolism of mRNA. Figure C shows the molecular function dot plot, indicating that ribonucleoprotein binding was the most significant and most frequent function of the randomly chosen 50-gene module set. Other genes include transporters of amino acids and peptidoglycans. Lysozyme activity had the third highest significance in the molecular function plot.

Figure D shows the cellular component analysis for the 50-gene module, where the highest gene frequency occurs within nuclear membrane processes, while the most significant genes appear to support intercellular bridging and azurophil granule lumens. Other notable functions that are not significant but frequently observed include endoplasmic reticulum lumens and endoplasmic reticulum protein-containing complexes. Other notable gene functions mostly refer to additional nuclear membrane processes, such as nuclear inclusion.

The morphology model presents an AUC of 85%, closely aligning with previous attempts using a new gene set.

## 5 Discussion

The convenience of the system is that, using preestablished prognostic labels such as the Moffitt classifiers, we can find gene sets that have similar expression trends that determine what label a case is. The model can be configured to only use a limited set of background genes to choose from, providing a means to explore a specific set of genes. Knowing that the labels are prognostic, research can be performed on gene sets and samples to understand prognosis. In summary, it saves time by automating the gene selection process. The disadvantage of this approach is that rerunning the model will not produce the same output twice. It is important to log all results beforehand. Interestingly, the best gene module selected two unmapped genes, which may be new genes or unmapped lncRNAs. The thresholds presented show high accuracy at 60 TPM and above, and the sensitivity and specificity of the modules are balanced.

The high-confidence classification label prediction task shows high performance, likely inhibited by occasional misnomers and inclusion of outlying cancer subtypes such as neuroendocrine cancers. The gene AUC was decently high for the second stage of the gene selection process, and the model converged across 30 epochs, showing no signs of overfitting. The important conclusion is that this new gene set is just one gene set produced from one run of the module, and many other gene networks with high predictive capacity can be used for molecular subtyping. This algorithm is limited by sample size and cohort variability.

The thresholding algorithm in the first stage of the model appears to have a high capability for filtering genes that are not as expressed as those in other modules, allowing modules that are consistently expressed across samples to have higher predictive power for classification. The same principle occurs with the Moffitt gene set, whereby markers are consistently highly expressed in classical and basal samples.

The high-confidence classification results demonstrate that morphology alone can approximate transcriptomic subtype structure when molecular ambiguity is minimized. Across five folds, the model consistently achieved test AUC values in the 0.85–0.93 range, indicating that the gene-constrained latent space captures biologically meaningful variation rather than superficial histologic correlates. Importantly, performance degradation in the low-confidence cohort suggests that the primary limitation is not architectural instability but transcriptomic ambiguity. Cases near the ssGSEA decision boundary likely represent molecular intermediates or heterogeneous tumors rather than clear subtype exemplars. This supports the interpretation that morphology reflects dominant molecular programs most clearly when the transcriptomic signal is strong, and that reduced performance in ambiguous cases reflects label noise or biological continuum rather than model collapse.

The hierarchical gene sampling results further indicate that predictive gene modules are sparse but non-random. As shown in the performance landscape of randomly sampled modules (Figure 3), most gene sets cluster around weak-signal regimes, while a small subset exhibits markedly elevated AUC. The recurrence of specific genes across high-performing modules suggests structured co-expression programs rather than stochastic selection effects. Moreover, the improvement from 200-gene to optimized 50-gene modules indicates that compression enhances signal-to-noise by removing weakly contributing genes. The graph-Laplacian regularization appears to constrain the morphology encoder toward biologically coherent latent representations, preventing reliance on staining artifacts or spurious texture cues. Together, these results support the feasibility of “virtual transcriptomics” as a constrained representation learning strategy rather than a purely predictive exercise.

## 6 Limitations

It should be noted that the training AUC maintained very high AUCs despite regularization and dropout, and that the performance on the high confidence samples can be limited by sample size and cohort variability. It is important to note that the low confidence samples contained a variety of cases, and there was class imbalance, where there was a 65-35 ratio between classical and basal cases in general. Furthermore there is a high number of low confidence classical cases in the low confidence cohort. While the model may struggle with low confidence cases, generally, low confidence cases are not considered clinically applicable or prognostic.

## 7 Conclusions

In conclusion, we developed a model that can select genes, profile them morphologically, and be used as a gene discovery program and a morphological subtyping application. In doing so, new genes that are prognostically significant can be indicated on biopsies and can be studied through transcriptomic analysis. This provides a means to explore new biomarkers. Importantly, the model demonstrates that morphology can approximate transcriptomic subtype structure when molecular signals are strong, while also revealing performance degradation in transcriptomically ambiguous cases. This distinction suggests that histologic phenotype reflects dominant molecular programs and that uncertainty in prediction may correspond to genuine biological intermediacy rather than model instability.

## 8 Ethics Statement

All datasets are anonymized. TCGA is publically available, PANCAN requires requests.

## 9 Author Contributions

A.L. conceived the study and was responsible for conceptualization, investigation, methodology, data preprocessing, gene ontology analysis, algorithm development, model implementation, validation, and testing. A.L. also wrote the original manuscript draft. A.A. curated the dataset and generated and prepared the embedding representations used in the study. M.K.K.N. supervised the project, acquired resources, provided critical review, and edited the manuscript. All authors reviewed and approved the final manuscript.

## 12 Data Availabilitye

Datasets and diagnostic slides are publically available and anonymized. The authors can not share PANCAN data, which can be requested here https://pancan.org/. The code is found here https://github.com/Alejandro21236/HasanovsModel. Here is the link to TCGA-BRCA, the openly available public dataset https://portal.gdc.cancer.gov/projects/TCGA-PAAD

